# The Effects of Yoga Nidra Practice on EEG Oscillations: A Systematic Review

**DOI:** 10.1101/2025.06.16.25329676

**Authors:** Vicky Kachera, Dimple Patel, Saurabh Mishra, Alka Mishra, James Gomes, Rahul Garg

## Abstract

**Background:** Yoga Nidra practice (YNP) is a systematic method of inducing physical, mental and emotional relaxation. Research suggests that YNP may enhance sleep quality and alleviate symptoms of anxiety and stress. Currently, research on YNP focuses on studying neural activity using EEG to determine the differences between YNP and sleep, but the findings remain fragmented. This review examines the relationship between YNP and EEG activity, based on quantitative studies reported in the literature.

**Methods:** Studies examining the effects of YNP on EEG oscillations, published in the English language, were searched in PubMed, Web of Science, and Scopus from inception to May 6, 2025. The quality of the studies was evaluated using the Mixed Methods Appraisal Tool (MMAT). Since a meta-analysis could not be done owing to heterogeneity in study designs, Power Spectral Density (PSD) and EEG-based sleep-related findings were synthesised narratively.

**Results:** Twelve studies with 326 participants met the inclusion criteria, employing varied designs including RCTs, as well as pre-post, within-subject and crossover studies. Only two studies met all the MMAT quality criteria. The most consistent finding was an increase in theta power among experienced practitioners, as reported in four studies. Changes in alpha, delta, beta, and gamma bands were inconsistent. A common observation was that participants with prior YNP experience remained awake during practice. Additionally, YNP was associated with improvements in sleep quality.

**Conclusion:** Yoga Nidra Practice shows promise in improving sleep-related outcomes and promoting deep relaxation. More well-designed studies with larger sample sizes and diverse populations are essential to better understand its impact on EEG and its underlying neural mechanisms.

**PROSPERO registration number:** CRD42024511747.

**Highlights:**

- Yoga Nidra practice (YNP) has significant effects on EEG oscillations.
- By facilitating deep relaxation, YNP improves sleep-related outcomes.
- No consistent patterns were observed in the Power Spectral Density of alpha, beta, delta and gamma bands.
- The findings elicit the need for further studies with larger sample sizes, diverse populations and longitudinal designs.

## 1. Introduction

The state of Yoga Nidra (YN) is an altered state of consciousness, not yet fully understood by modern science (1). According to Parker et al. (2013), the YN state resembles deep non-rapid eye movement (REM) sleep, while awareness remains unbroken (1). Although no published scientific studies have verified this phenomenon, pilot reports documented it in accomplished yogis such as Swami Rama and his disciple Swami Veda Bharati, who had the ability to consciously enter the YN state (2,3). Yoga Nidra practice (YNP) is an ancient technique rooted in the Tantras (4) and defined as a systematic method for achieving complete physical, mental, and emotional relaxation, and was revived in modern times by yogis such as Swami Satyananda Saraswati and Swami Rama, who emphasised that the Yoga Nidra state can be attained through sustained practice (3,4).

Since YNP has its roots in ancient Indian traditions, there are many variations in the practice. Some of these variants include the ‘61-Points Relaxation Exercise’ developed by Swami Rama of the Himalayan yoga tradition (5), Marmanasthanam Kriya, from the Rishiculture Ashtanga Yoga (Gitananda Tradition) (6), and Integrative Restoration (iRest), a modern adaptation developed by Richard Miller based on the ancient YNP (7). Dr. Andrew Huberman considers YN as a form of Non-Sleep Deep Rest (NSDR), a term he coined to describe mindfulness practices that induce deep relaxation without leading to sleep (8,9). It has been reported that YNP improves mental health by alleviating symptoms of stress, anxiety, and depression (10). Several clinical trials employing YNP have shown improvements in sleep quality and reductions in symptoms of insomnia (11–14). Other studies have shown that YNP lowers blood pressure in hypertensive patients (15) and improves the psychological well-being of women experiencing menstrual disorders (16).

Parker et al. (2013) suggested a definition of YNP that describes it electrophysiologically across several levels (1). In the initial stages of practice, individuals typically experience deep relaxation marked by increased alpha wave activity, which may gradually shift toward theta waves with deeper engagement (1). Swami Satyananda Saraswati also emphasised the predominance of alpha waves during YNP, associating it with profound mental, emotional, and muscular relaxation (4). Both Parker et al. (2013) and Swami Satyananda proposed that in the deeper YN state, practitioners may consciously enter a state characterised by delta wave activity, typically associated with deep, non-REM sleep, while remaining fully aware of the environment (1,4). Building on these theoretical perspectives and published literature, recent studies have employed EEG to objectively examine the brain wave patterns during YNP. However, drawing definitive conclusions about specific brainwave patterns remains challenging owing to methodological heterogeneity resulting from variability in YN protocols, practitioner experience, and study designs. Additionally, some studies have recorded EEG activity during practice, while others have assessed post-practice effects. To date, no comprehensive review has systematically analysed EEG findings across YNP studies. Therefore, the objective of this systematic review is to identify and characterise studies examining brain wave activity associated with YNP.

## 2. Methods

This systematic review adhered to the updated 2020 Preferred Reporting Items for Systematic Reviews and Meta-Analyses (PRISMA) guidelines (17), and the protocol was registered in PROSPERO (CRD42024511747).

### 2.1 Information Sources and Search Strategy

A PROSPERO search conducted on January 7, 2024, revealed no registered systematic reviews focusing specifically on YN and its effects on brain waves using EEG. Three databases namely PubMed, Web of Science, and Scopus were searched for articles published from the inception of each database until May 6, 2025, using broad search terms such as ‘Yoga Nidra,’ ‘Yoganidra,’ ‘Yogic Sleep,’ ‘Psychic Sleep,’ ‘Hypnagogic Sleep,’ ‘NSDR,’ ‘iRest,’ ‘Electroencephalogram,’ ‘EEG,’ and ‘Brain Waves.’ These terms were intentionally chosen to capture diverse terminology associated with YN and related neurophysiological states. Terms such as ‘Yoganidra,’ ‘Yogic Sleep,’ ‘Psychic Sleep,’ and ‘Hypnagogic Sleep’ addressed conceptual variations, while ‘EEG,’ ‘Electroencephalogram,’ and ‘Brain Waves’ ensured inclusion of studies involving electrophysiological measurements. No filters or restrictions were applied for the search process. The detailed search query for all databases has been provided in Supplementary Table A.

### 2.2 Study Selection Process

The detailed eligibility criteria are summarised in Table 1. Findings from each database were imported into RAYYAN for deduplication and screening (18). Two reviewers (VK and DP) independently screened the articles by title and abstract according to the predetermined inclusion and exclusion criteria. Full-text articles were downloaded and thoroughly reviewed to confirm their eligibility. Additionally, the reference lists of the selected articles were examined to identify any other relevant studies. Before the final inclusion of articles, consensus was reached, and any disagreements were resolved through discussion. Study authors were contacted by email to clarify doubts, confirm details, or request additional information as needed. Referencing was managed using Zotero 6.0.36, an open-access reference management tool (19).

**Table 1:**
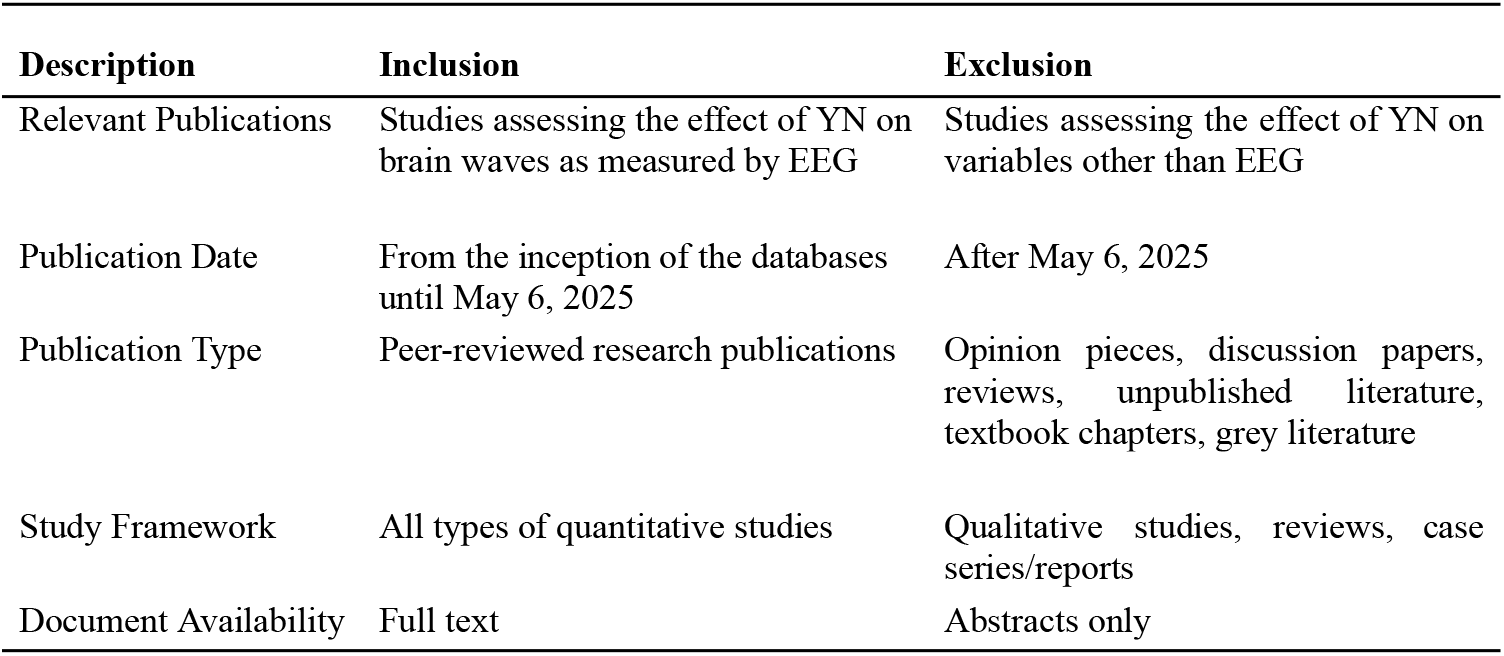
Inclusion and Exclusion Criteria for Study Selection.

| **Description** | **Inclusion** | **Exclusion** |
| --- | --- | --- |
| Relevant Publications | Studies assessing the effect of YN on brain waves as measured by EEG | Studies assessing the effect of YN on variables other than EEG |
| Publication Date | From the inception of the databases until May 6, 2025 | After May 6, 2025 |
| Publication Type | Peer-reviewed research publications | Opinion pieces, discussion papers, reviews, unpublished literature, textbook chapters, grey literature |
| Study Framework | All types of quantitative studies | Qualitative studies, reviews, case series/reports |
| Document Availability | Full text | Abstracts only |

### 2.3 Data Collection Process and Data Items

The data collection process involved creating a table, with the reviewer VK extracting the data including the following aspects: study information (title, journal, author(s), year, country, study design, blinding, randomisation details); sample characteristics (inclusion-exclusion criteria, sample size (n), gender, age range); control and intervention details (control/comparator group details, experimental intervention details, duration and frequency); EEG data and methodology (EEG channels, locations analysed, impedance, sampling rate, feature extraction, EEG methodology); and key findings (EEG). This data was verified by another reviewer (DP), with any disagreements resolved by a third reviewer (RG). Data extraction was performed manually, without the use of automation tools.

### 2.4 Quality Assessment

The MMAT tool can appraise heterogeneous empirical studies across five categories: Qualitative, Quantitative Randomised Controlled Trials (RCTs), Quantitative Non-Randomised, Quantitative Descriptive, and Mixed Methods studies (20). It has been implemented in various mixed-method systematic reviews (21–23). Two reviewers (VK and DP) independently evaluated the quality of all included studies by using the 2018 version of MMAT (20,24). Each question in the selected category was rated as ‘Yes,’ ‘No,’ or ‘Can’t tell’ (when sufficient information was not provided to answer ‘Yes’ or ‘No’) (20). Any discrepancies in ratings were resolved through discussion, and a third independent reviewer (RG) was involved whenever needed. For ‘Can’t tell’ ratings, the corresponding authors of the studies were contacted by email for more information or clarification. The results are presented as follows: a score of 100% if all five quality criteria are met, 80% if four, 60% if three, 40% if two, and 20% if one (25). No automation or AI-based tools were used to assess study quality.

### 2.5 Statistical Synthesis

The heterogeneity in the YNP type, duration, results, population, and study design did not permit a meta-analysis. Instead, the results were reported in tabular form and summarised narratively.

## 3. Results

### 3.1 Search Results

A total of 36 records were identified through our search strategy, including 10 from PubMed, seven from Web of Science, 16 from Scopus, and three from reference searches. Fifteen duplicates were removed using RAYYAN, leaving 21 articles. After screening titles and abstracts in RAYYAN, 14 articles remained. One article’s (26) full text was unavailable, and another was published in a journal not indexed in global databases, leaving 12 articles for quality appraisal. See Figure 1 for the PRISMA 2020 flow diagram.

**Figure 1:**
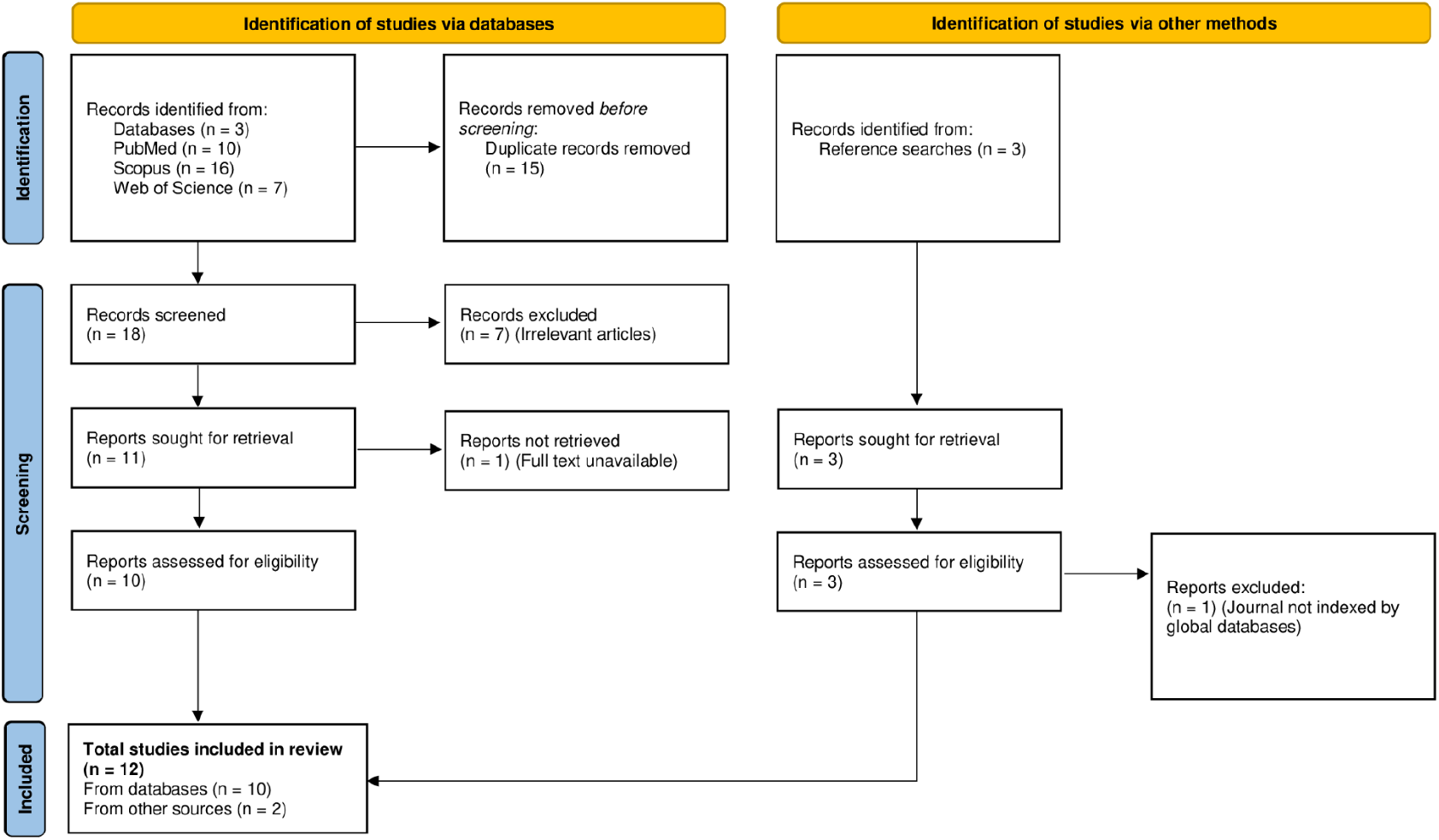
PRISMA flow diagram

### 3.2 Study and Participant Characteristics

In Tables 2a and 2b, the characteristics of the included studies have been summarised. The review includes 12 papers that met the eligibility criteria: Three were pre-post interventions (14,27,28), three RCTs (29–31), one within-subject randomised crossover study (32), three within-subject studies (33–35), and two were pre-post with a control group (36,37). Most studies (n = 8) were published within the last 10 years. From these 12 papers, data for a pool of 326 participants were curated: Three papers had 50 to 60 participants, four had 20 to 49, and five had fewer than 20. Three studies involved only male participants, one involved only female participants, while three included both male and female participants. Two studies had an equal number of male and female participants, another two did not report gender information, while one mentioned both genders were involved but did not provide specific numbers. The age of participants ranged from 18 to 90 years. Two studies included participants with yoga experience ranging from 5 to 26 years (32,33), while one focused on long-term YN practitioners (35). Two studies involved healthy novices (14,27), and one involved novice insomnia patients (30). The remaining six studies did not report participants’ yoga experience.

**Table 2a:** Characteristics of Studies Employing Power Spectral Density (PSD) Analysis of EEG During Yoga Nidra Practice.

| Subject Characteristics |  |  |  |
| --- | --- | --- | --- |
| Study Design<br>(Country) | Description and<br>Sample Size (N) | Gender (M/F)<br>Age Mean (yrs)<br>Age Range (yrs) | Reference |
| Pre-post intervention<br>(India) | Healthy novices<br>N = 30 | M/F: (15/15)<br>Mean: 35.53 ± 8.69 | (14) |
| Within-subject,<br>randomised crossover<br>(UK) | Meditation teachers with<br>7 to 26 years of<br>experience.<br>N = 8 | M/F (8/0)<br>Range 31–50 | (32) |
| Within-subjects (NR) | Yoga practitioners with<br>over 5 years of<br>experience in Kriya<br>Yoga, YNP, and other<br>yoga techniques.<br>N = 9 | M/F (6/3)<br>Range: 23–41 | (33) |
| RCT (USA) | Novices with insomnia. |  | (30) |
|  | YNP (N= 9) | M/F (3/6)<br>Mean: 33 ± 7 |  |
|  | CG (N= 9) | M/F(1/8)<br>Mean:30 ± 6 |  |
| Within-subjects (France) | Healthy practitioners<br>with 10 to 12 years of<br>formal YNP.<br>N = 6 | M/F (2/4)<br>Range: 31–74 | (35) |

RCT: Randomised controlled trial, CG: Control group, NR: Not reported

**Table 2b:** EEG Outcomes: Following YNP or Interventions Combined with YNP.

| Study Design<br>(Country) | Subject Characteristics |  | Reference |
| --- | --- | --- | --- |
|  | Description and<br>Sample Size (N) | Gender (M/F)<br>Age Mean (yrs)<br>Age Range (yrs) |  |
| RCT (India) | Athletes |  | (29) |
|  | YNP (N= 28) | Only Males<br>Mean: 22.45 ± 4.53 |  |
|  | CG (N= 29) |  |  |
| RCT (India) | Migraine patients |  | (31) |
|  | YNP (N= 30) | M/F<br>Range: 18–25 |  |
|  | CG (N= 30) |  |  |
| Pre-post interventional<br>cohort (India) | Healthy novice<br>N= 41 | Only Males<br>Mean: 25.146 ± 2.275 | (27) |
| Within-subjects (USA) | Interested individuals<br>N = 10 | M/F: (5/5)<br>Mean: 41.2<br>Range: 26-90 | (34) |
| Studies that included Yoga Nidra in combination with other interventions as the independent variable |  |  |  |
| Pre-Post with control<br>group (India) | Working women |  | (36) |
|  | YNP (N= 25) | Only Females<br>Range: 25-39 |  |
|  | CG (N= 25) |  |  |
| Pre-Post with control<br>group (India) | Physically fit students |  | (37) |
|  | YNP (N= 20) | NR |  |
|  | CG (N= 20) |  |  |
| Pre-Post (India) | College students.<br>N = 40 | Range: 18-25 | (28) |

### 3.4 Results of Quality Assessment

Table 3 summarises the quality assessment results. The quality assessment provides critical insight into the methodological rigour and reliability of the included studies. Of the 12 studies, two met 100% of the criteria (14,30), six met 80% (27,29,31,33,34,36), one met 60% (35), two met 40% (32,37), and one met 20% (28). Although none of the studies were excluded based on quality, the variability in scores highlights the importance of considering study quality when drawing conclusions and making recommendations (20).

**Table 3:** Assessment of Study Quality Following the Mixed Methods Appraisal Tool (MMAT) (20)

| **1. RANDOMISED CONTROLLED TRIALS** |  |  |  |  |  |  |
| --- | --- | --- | --- | --- | --- | --- |
| **1. Is randomisation appropriately performed?** | **2. Are the groups comparable at baseline?** | **3. Are there complete outcome data?** | **4. Are outcome assessors blinded to the intervention provided?** | **5. Did the participants adhere to the assigned intervention?** | **Quality (%)** | **Reference** |
| Yes | Yes | No | Yes | Yes | 80 | (29) |
| Can't tell | Yes | No | Can't tell | Yes | 40 | (32) |
| Yes | Yes | Yes | Yes | Yes | 100 | (30) |
| Yes | Yes | Yes | Can't tell | Yes | 80 | (31) |
| **2. NON-RANDOMISED STUDIES** |  |  |  |  |  |  |
| **1. Are the participants representative of the target population?** | **2. Are measurements appropriate regarding both the outcome and intervention (or exposure)?** | **3. Are there complete outcome data?** | **4. Are the confounders accounted for in the design and analysis?** | **5. During the study period, is the intervention administered (or exposure occurred) as intended?** | **Quality (%)** | **Reference** |
| Yes | Yes | Can't tell | Can't tell | Can't tell | 40 | (37) |
| Yes | Yes | Yes | Can't tell | Yes | 80 | (36) |
| Yes | Yes | Yes | Yes | Yes | 100 | (14) |
| Yes | Yes | Yes | No | Yes | 80 | (27) |
| Yes | Can't tell | Can't tell | Can't tell | Can't tell | 20 | (28) |
| Yes | Yes | Yes | Can't tell | Yes | 80 | (33) |
| Yes | Yes | Yes | No | Yes | 80 | (34) |
| Yes | Yes | No | No | Yes | 60 | (35) |

### 3.5 Intervention Characteristics

A detailed overview of EEG acquisition parameters, including electrode locations, sampling rates, and feature extraction methods, is presented in Table 4, while the characteristics of the interventions are provided in Tables 5 and 6. Two studies used Swami Satyananda Saraswati’s YNP developed for insomnia (14,30), while the other 10 did not specify the tradition. The session durations ranged from 20 minutes (27) to 2 hours (35), with the most common duration being 27-30 minutes (14,28–30,36). Two studies did not report the duration of YN sessions (31,37). The duration of intervention was largely variable among the studies. Four studies evaluated the effect of a single session of YNP (32–34,30); while others ranged from 12 sessions over 6 days (35) to daily sessions for 45 days (36). One study did not specify the exact intervention period, only stating it was between 3 and 6 months (37). All studies focused on YN as an independent variable except for three (28,36,37), which combined YNP with other yogic practices such as Nadi Shodhan Pranayama, Pranakarshan Pranayama, Nine-Centre Meditation, and Pragya Yoga.

**Table 4:** Technical Specifications of EEG Recording and Analysis in Yoga Nidra Practice Studies.

| EEG Channels | Locations Analysed | Impedance (kΩ) | Sampling Rate (HZ) | Feature Extraction | Reference |
| --- | --- | --- | --- | --- | --- |
| 19 | C3, C4, Cz, F3, F4, F7, F8, Fz, Fp1, Fp2, P3, P4, Pz, T3, T4, T5, T6, and O1, O2. | $\leq 5$ | 500 | PSD | (14) |
| 3 | F3, O1, and C3. | NR | 256 | PSD | (27) |
| NR | Electrodes were positioned at O1, O2, F3, C3, C4, and F4, along with EOG and EMG sites. | $< 5$ | NR | PSG analysis | (27) |
| NR | O1-M2/O2-M1, C3-M2/C4-M1, F3-M2/F4-M1, and two EOG and EMG. | NR | NR | NR | (29) |
| 16 | NR | NR | NR | PSD | (32) |
| 16 | NR | NR | NR | PSD | (33) |
| 128 | As per standard 10-20 system with EOG | $\leq 50$ | 600 | FFT and ICA | (34) |
| NR | F3, O1, C3, EOG and chin locations. | NR | NR | PSD, PSG sleep scoring | (30) |
| NR | NR | NR | NR | NR | (31) |
| 128 | NR | $< 50$ | 500 | PSD | (35) |
| NR | NR | NR | NR | NR | (36) |
| 8 | O1, O2, F3, F4, P3, P4, and C3, C4. | NR | 256 | DWT | (37) |
| NR | NR | NR | NR | NR | (28) |

O: Occipital, F: Frontal, T: Temporal, C: Central, DWT: Discrete wavelet transform, FFT: Fast fourier transformation, ICA: Independent component analysis, PSG: Polysomnographic, NR: Not reported

**Table 5:** Power Spectral Density (PSD) Changes in EEG During Yoga Nidra Practice.

| Experimental Protocol | Control Group | Baseline Condition Description | Comparison Condition | Findings Across Frequency Bands |  |  |  |  | Remarks | Reference |
| --- | --- | --- | --- | --- | --- | --- | --- | --- | --- | --- |
| | | | | $\Delta$ Delta | $\Delta$ Theta | $\Delta$ Alpha | $\Delta$ Beta | $\Delta$ Gamma | | |
| SSS's YNP for insomnia was practised for 2 weeks with ~27-min audio; EEG was recorded during YNP after 2 weeks. | NA | Four cycles of 1-min eyes closed and 1 min eyes open (8 min total). | Baseline | Central ↑<br>Prefrontal ↓ | No change in the central region. | Occipital and parietal ↓ | NR | NR | PSG data confirmed the awake state of participants. | (14) |
| Single 72-minute YNP session. EEG was recorded during YNP for both groups. | Yes | NA | Listened to speech (same voice as YNP), eyes closed, 72 min. | NR | ↑ 9% (NS) | ↓ 8% (NS) | NR | NR | - | (32) |
| Two-hour Tantric Kriya Yoga (preparation), followed by a single 45-minute YNP session. EEG was recorded during the entire YNP session. | NA | Supine position, no sensory input, eyes covered, pre-YNP. | Baseline | NR | Increased | ↓ 2% (NS) | NR | NR | Alpha was similar to the resting state, indicating wakefulness. | (33) |
| SSS's YNP for insomnia was used. Single 30-minute YNP session followed by 60-minute quiet rest (nap) at V2. EEG was recorded throughout. | Yes | Supine position, no sensory input, 90-min quiet rest at visit 1(baseline). EEG was recorded during visit 1. | Baseline and 90-minute quiet rest at V2 (CG). | O1: ↓ 32% in the CG and ↓ 17% in the YNP group between V1 and V2 (NS). | During the nap, ↓ less in the YNP group compared to the CG (NS). | Slight ↑ at O1 in the YNP group and slight ↓ in the CG from V1 to V2 (NS within & between). | NR | NR | 89% of the intervention group slept, compared to 78% of controls. | (30) |
| Twelve group YNP sessions over 6 days; each 2 hours. EEG was recorded during each YNP session. | NA | Supine position, spontaneous breathing, 10 min, pre-YNP. | Baseline | No change | Widespread ↑ during mental visualisation, ↓ later. | Initial widespread ↑, then ↓. | Initial widespread ↑, then ↓. | Widespread gamma ↑ in late stages; | No K-complexes or sleep spindles observed. | (35) |
| Single 65-min YNP session performed via audio seated with eyes closed; EEG recorded throughout. | NA | NR | NR | Rhythmic waves below the infraslow range (about 8-30 min) and a near-DC component, primarily in the frontal midline and inferior hemispheric regions.<sup>1</sup> |  |  |  |  |  | (34) |

NA: Not available, NR: Not reported, Sign.: Significant, NS: Not significant SSS: Swami Satyananda Saraswati, ↑: Increase, ↓: Decrease, CG: Control group, V1: visit 1, V2: visit 2
<sup>1</sup>Rodin et al., 2017 recorded EEG during YNP, but did not report PSD analysis.

**Table 6:** EEG Outcomes: Following YNP or Interventions Combined with YNP.

| Experimental Protocol | Control Group | Comparison Condition | Key Findings (EEG) | Reference |
| --- | --- | --- | --- | --- |
| YNP (20 min/day) for 4 weeks; overnight PSG recorded at baseline and after 4 weeks. EEG was recorded during daytime cognitive tasks. | NA | Baseline | Significant improvements in wake-after-sleep onset, sleep efficiency, and delta (%) in slow-wave sleep. | (27) |
|  |  | Control with Eyes Open (CEO) and Control with Eyes Closed (CEC) | O1: ↑ theta during VOLT, ↑ delta/beta during BART (after 2 weeks of YN)<br>C3: ↑ theta during VOLT and MPT (after 1 week of YN)<br>F3: ↑ delta/alpha, ↑ delta/theta during ERT (after 1 week of YN) |  |
| YNP (~27.2 min/day) for 4 weeks, with one pre-sleep hygiene session; overnight EEG recorded at baseline and after 2 weeks. | PMR practised before sleep (10–15 min) for 4 weeks, preceded by a sleep hygiene session. | Baseline | Significant improvements in sleep onset latency and time in bed in both groups. | (29) |
| YNP for 6 weeks. EEG was recorded at baseline and after 6 weeks. | Received supine rest for 6 weeks. | Control group | A significant increase in “Alpha frequency” in the YNP group; no significant changes in the other EEG waveforms. | (31) |
| Pragya Yoga, NSP, and YNP (30 min) practised together (60 min/day) for 45 days; Alpha EEG biofeedback was recorded at baseline and after 45 days. | NR | Control group | A significant increase in “alpha EEG” was observed in the intervention group. | (36) |
| YNP, NSP, and NCM were first practised as preparation, then individually (the duration was not explicitly reported). EEG was recorded for 40 min at baseline and after 6 months. | NR | Control group | “Alpha and beta frequencies” decreased in the YNP group. | (37) |
| YNP (30 min) and PP (15 min) practised daily for 40 days; Alpha EEG biofeedback was recorded at baseline and after 40 days. | NA | Baseline | A significant increase in “alpha EEG”. | (28) |

O1: Occipital region, C3: Central region VOLT: Visual Object Learning Task, MPT: Motor Praxis Task, BART: Balloon Analog Risk Task, ERT: Emotion Recognition Task, ↑: Increase, ↓: Decrease, PMR: Progressive Muscle Relaxation, NSP: Nadi Shodhan Pranayama, PP: Pranakarshan Pranayama, NCM: Nine-Centre Meditation

### 3.6 EEG Power Analysis During Yoga Nidra Practice

Table 5 provides a detailed overview of power spectral density (PSD) changes observed during YNP across studies. All studies reported PSD analysis during YNP except the study by Rodin et al. (2017), which reported “near-DC activity that extended beyond one hour, as well as rhythmic wave durations ranging from about 10 to 35 min” during a single 65-minute YNP session” primarily in the frontal midline and inferior hemispheric regions, suggesting a deeply altered neural state (34). The following subsections provide a breakdown of results by frequency band.

#### 3.6.1 Delta Band

Delta activity showed varied changes across studies. Datta et al. (2022) reported an increase in delta power in the central region and a decrease in the prefrontal region during YNP compared to the pre-YNP baseline (14). In another study, delta power decreased by 32% in the control group and by 17% in the YNP group; however, these changes were not statistically significant (30). Zaccaro et al. (2020) found no significant change in delta power during the YNP session (35).

#### 3.6.2 Theta Band

Changes in theta power during YNP have shown a general trend toward an increase, although most findings did not reach statistical significance. Kjaer et al. (2002) observed a 9% increase (non-significant) in theta power during YNP compared to the control condition (32), while Lou et al. (1999) reported an 11% increase during YNP compared to a pre-YNP resting state (33). Zaccaro et al. (2021) found that theta power increased during the mental visualisation stage of YNP but gradually declined thereafter (35). Sharpe et al. (2023) noted that theta power decreased less in the YNP group than in the control group during a post-YNP nap session; however, this difference was not statistically significant (30).

#### 3.6.3 Alpha Band

Changes in alpha power during YNP varied across studies, with differing patterns observed depending on participant characteristics and session conditions. Kjaer et al. (2002) reported an 8% decrease in alpha power compared to the control condition (32), and Datta et al. (2022) similarly found reduced alpha power in novices, though PSG (Polysomnographic) analysis confirmed participants remained awake (14). In contrast, Lou et al. (1999) reported only a 2% non-significant decrease from resting baseline, suggesting wakefulness during YNP (33). Zaccaro et al. (2021) observed an initial increase in alpha power that gradually declined over time (35). Sharpe et al. (2023) found a slight increase at O1 in the YNP group and a decrease in the control group; however, PSG analysis showed 89% of participants fell asleep during the session (30). Additionally, post-YNP nap data showed a smaller alpha power decrease in the YNP group than in controls, though the difference was not statistically significant (30).

#### 3.6.4 Beta and Gamma Bands

Changes in beta and gamma power were reported only by Zaccaro et al. (2021) (35). Beta power initially increased during YNP and gradually declined as the session progressed. Gamma power showed an increase during the final stages of YNP (35).

### 3.7 EEG-Based Sleep Outcomes

As summarised in Table 6, two studies assessed the impact of YNP on objective sleep parameters using overnight EEG recordings. Datta et al. (2023) reported significant improvements in wake-after-sleep onset, sleep efficiency, and increased delta percentage during slow-wave sleep after four weeks of daily YN practice (27). Similarly, Datta, Yadav et al. (2022) found significant improvements in sleep onset latency and time in bed following YNP, with comparable improvements also observed following progressive muscle relaxation in the control group (29). These findings suggest that YNP may enhance sleep continuity and quality, as evidenced by changes in electrophysiological markers of sleep.

### 3.8 EEG Power Analysis Following Yoga Nidra Practice or Combined Interventions

As detailed in Table 6, several studies examined power spectral density (PSD) changes following YNP, either as a standalone intervention or in combination with other yogic techniques. Datta et al. (2023) reported a significant increase in delta and theta power at the occipital site (O1) during a cognitive task after two weeks of YNP (27). Shashikiran et al. (2022) observed a significant increase in the alpha power spectrum in migraine patients following six weeks of YNP, compared to a control group receiving supine rest, with no significant changes in other frequency power bands (31). Other studies reported changes in the power of frequency bands following YNP combined with additional yogic practices. Bharadwaj et al. (2013) found a significant increase in “alpha activity” after a 45-day intervention incorporating Pragya Yoga, Nadi Shodhan Pranayama, and YNP among working women (36). Kumar and Joshi (2009) also reported increased “alpha activity” after 40 days of combined YNP and Pranakarshan Pranayama practice in college students (28). In contrast, Anuradha et al. (2022) observed reductions in both “alpha and beta frequencies” following six months of YNP combined with Nine-Centre Meditation and Nadi Shodhan Pranayama among students (37). These findings suggest that YNP, especially when practised alongside other techniques, may influence alpha and beta band powers, although the direction and extent of change appear to depend on the specific intervention protocol and participant characteristics. Overall, due to poor reporting, low methodological quality, and inconsistencies in the results related to alpha power, no definitive conclusions can be drawn.

## 4. Discussion

This systematic review summarises findings from 12 studies involving 326 participants investigating the effects of Yoga Nidra practice (YNP) on EEG activity. As shown in Tables 2a and 2b, variation in participant experience with YNP and participant characteristics should be considered when interpreting the results. The included studies span diverse populations, including long-term practitioners, healthy novices, and individuals with conditions such as insomnia and migraine. EEG outcomes during and following YNP are summarised in Table 5 (PSD changes during YNP) and Table 6 (other post-intervention EEG measures).

According to the electrophysiological definition of Yoga Nidra Practice (YNP) proposed by Parker et al. (2013) (1), it involves a progressive shift in brain activity: an initial increase in alpha power with theta becoming more prominent as the practice deepens. This definition forms the basis for interpreting EEG changes observed during YNP. However, the current body of evidence shows inconsistent findings in relation to this theoretical hypothesis, particularly pertaining to alpha power. For example, Datta et al. (2022) (14) reported a significant decrease in occipital and parietal alpha power in novice insomnia patients, compared to baseline, after a 27-minute YNP session, which contradicts the expected early alpha enhancement. Similarly, Kjaer et al. (2002) (32) and Lou et al. (1999) (33) found non-significant decreases in alpha power. Kjaer et al. (2002) (32) reported this when compared to an active control group, while Lou et al. (1999) (33) observed it relative to baseline. Both studies involved experienced practitioners but varied in session length (72 minutes versus 45 minutes) and comparison conditions, making interpretation difficult. In contrast, Sharpe et al. (2023) (30), using a 30-minute protocol with insomnia patients, reported a non-significant increase in alpha power at the O1 site (Table 5), which loosely supports the early-stage alpha increase described by Parker et al. (2013) (1) hypothesis.

The study by Zaccaro et al. (2021) (35) demonstrates stronger alignment with the definition given by Parker et al. (2013) (1). Although limited to a single experienced subject, this study (35) conducted a stage-wise analysis, revealing an initial widespread alpha increase followed by a subsequent decrease as the session progressed. During the mental visualisation stage, theta power increased, then declined later (35). This temporal EEG pattern (35) closely mirrors the progression described by Parker et al. (2013) (1) and suggests that stage-specific analysis with a large sample size may be crucial for capturing the dynamic nature of YNP-induced EEG changes.

Theta power findings across studies were somewhat more consistent with theoretical expectations. Lou et al. (1999) (33), with a sample of nine experienced practitioners, reported a significant increase in theta power compared to baseline. Notably, this group (33) engaged in a two-hour Tantric Kriya Yoga practice prior to the YNP session, which may have deepened their meditative state and contributed to the pronounced theta activity. Kjaer et al. (2002) (32), using a similarly small sample of eight, observed a non-significant increase relative to an active control group, possibly due to limited statistical power. Despite these small samples, both studies (32,33) support the notion that theta becomes more prominent in the deeper stages of YNP, particularly among experienced practitioners. In contrast, Datta et al. (2022) (14) reported no significant change in theta activity in novices, suggesting that experience level, preparatory practices, and practice depth may be critical for the emergence of theta-dominant states (Table 5).

Apart from this, a few studies examined sleep-related EEG outcomes following regular YNP. These findings, though limited, suggest improvements such as enhanced sleep efficiency, reduced wake-after-sleep onset, increased delta power during slow-wave sleep, shortened sleep onset latency, and extended total time in bed (27,29). Further, these results imply that YNP may influence broader neurophysiological processes associated with restorative sleep. Nevertheless, due to heterogeneity in YNP protocols, small sample sizes, lack of stage-wise EEG reporting, and varying control conditions, it is currently not possible to draw firm conclusions about consistent EEG patterns during YNP. While elements of Parker et al. (2013) (1) hypothesis, particularly theta enhancement in deeper stages, are supported in two studies, the overall evidence remains mixed.

To clarify the neurophysiological mechanisms of YNP and better test the hypothesis proposed by Parker et al. (2013) (1), future research should prioritise larger sample sizes, incorporate both baseline and active control comparisons, and conduct stage-specific and region-wise EEG analyses. Detailed reporting of EEG acquisition parameters (for example, sampling rates and preprocessing methods) and clear documentation of the YNP protocol used, including the lineage and version, are essential to reduce variability. Ideally, researchers should adopt a standardised protocol, such as the basic version of Yoga Nidra developed by Swami Satyananda Saraswati, to enhance consistency across studies. Furthermore, transparency through preregistration and open data sharing will support reproducibility and comparability.

## 5. Conclusion

This review suggests that Yoga Nidra may modulate alpha and theta power, with some evidence of stage-specific effects in experienced practitioners. Additionally, two studies reported improvements in sleep-related outcomes, indicating potential benefits beyond the immediate practice. However, inconsistent findings and methodological variation limit definitive conclusions.

## Data Availability

All data produced in the present work are contained in the manuscript.

## Credit Authorship Contribution Statement

Rahul Garg: Conceptualisation, Supervision, Writing – review and editing. Vicky Kachera: Data curation, Formal analysis, Writing – original draft, Writing – review and editing. Dimple Patel: Data curation, Formal analysis, Writing – review and editing. Alka Mishra: Writing – Final review and editing. Saurabh Mishra: Writing – Final review and editing. James Gomes: Writing – Final review and editing.

## Declaration of Interests

The authors declare that there are no competing financial or personal relationships that could inappropriately influence the work reported in this paper.

## Funding Sources

This research did not receive any specific grant from funding agencies in the public, commercial, or not-for-profit sectors.

## Footnotes

1 Rodin et al., 2017 recorded EEG during YNP, but did not report PSD analysis.

## Notes

### Competing Interest Statement

The authors have declared no competing interest.

### Funding Statement

This study did not receive any funding.

### Summary of Updates

1. Table formatting has been improved for better readability. 2. Minor grammatical errors have been corrected to enhance clarity. 3. The discussion section has been revised. In the previous version, we highlighted consistent findings on theta power based primarily on mostly non-significant results. The updated discussion now places greater emphasis on recommendations for future studies. 4. Minor revisions have also been made to the Author Contributions section.

