## Supplementary Table A for "The Effects of Yoga Nidra Practice on EEG Oscillations: A Systematic Review"

Table A: Search query summary

| Database | Search query | |
| --- | --- | --- |
| PubMed | #1 | "Yoga Nidra" [Title/Abstract] OR "Yoganidra" [Title/Abstract] OR "Yoga-nidra" [Title/Abstract] OR "Yogic Sleep" [Title/Abstract] OR "Psychic Sleep" [Title/Abstract] OR "Hypnagogic Sleep" [Title/Abstract] OR "iRest" [Title/Abstract] OR "NSDR" [Title/Abstract] |
|  | #2 | "EEG" [Title/Abstract] OR "Electroencephalogram" [Title/Abstract] OR "Brain Waves" [Title/Abstract] |
|  | #3 | #1 AND #2 |
| Web of Science | 1. | (((((((TI=("Yoga Nidra")) OR TI=("Yoganidra")) OR TI=("Yoga-nidra")) OR TI=("Yogic Sleep")) OR TI=("Psychic Sleep")) OR TI=("Hypnagogic Sleep")) OR TI=("NSDR")) OR TI=("iRest") |
|  | 2. | (((((((AB=("Yoga Nidra")) OR AB=("Yoganidra")) OR AB=("Yoga-nidra")) OR AB=("Yogic Sleep")) OR AB=("Psychic Sleep")) OR AB=("Hypnagogic Sleep")) OR AB=("NSDR")) OR AB=("iRest") |
|  | 3. | ((TI=("EEG")) OR TI=("Electroencephalogram")) OR TI=(Brain Wave*) |
|  | 4. | ((AB=("EEG")) OR AB=("Electroencephalogram")) OR AB=(Brain Wave*) |
|  | 5. | #1 OR #2 |
|  | 6. | #3 OR #4 |
|  | 7. | #5 AND #6 |
| Scopus | 1. | ( TITLE-ABS-KEY ( "Yoga Nidra" ) OR TITLE-ABS-KEY ( "Yoganidra" ) OR TITLE-ABS-KEY ( "Yoga-Nidra" ) OR TITLE-ABS-KEY ( "Yogic Sleep" ) OR TITLE-ABS-KEY ( "Psychic Sleep" ) OR TITLE-ABS-KEY ( "Hypnagogic Sleep" ) OR TITLE-ABS-KEY ( "NSDR" ) OR TITLE-ABS-KEY ( "iRest" ) AND TITLE-ABS-KEY ( "EEG" ) OR TITLE-ABS-KEY ( "Electroencephalogram" ) OR TITLE-ABS-KEY ( "Brain Waves" ) ) |

Note: Inclusion and exclusion criteria were slightly modified post-registration on PROSPERO. Qualitative and observational studies were excluded as they did not align with the objective of this review; none of the studies were excluded based on their MMAT scores, but the quality percentages have been mentioned.
